# Effectiveness Of Telephone Instructions In Performing Cardiopulmonary Resuscitation: A Systematic Review

**DOI:** 10.1101/2023.06.24.23291852

**Authors:** Marios Charalampopoulos, Panagiota Triantafyllaki, Christina-Athanasia Sampani, Christos Triantafyllou, Dimitrios Papageorgiou

**Author notes:** **Corresponding author:** Christos Triantafyllou, Papadiamantopoulou 123, Goudi Athens, PC 11527. On Behalf: ICU Follow-Up Care Lab, Department of Nursing, University of West Attica, Athens, Greece.

## Abstract

**Introduction:** Cardiopulmonary resuscitation (CPR) refers to the emergency procedure that must be applied to any person suffering from cardiopulmonary arrest to keep them alive. Although CPR is an urgent and critical procedure, it consists of specific steps and skills, which must be applied in a specific sequence and with the best possible quality in order to be effective. Due to this importance, some National First Aid Centers around the world provide the necessary instructions to the potential rescuer by telephone (Telecommunicator CPR - TCPR), in order to ensure the proper appliance and effectiveness.

**Aim:** To gather scientific information from the existing surveys regarding the effectiveness of telephone instructions in performing cardiopulmonary resuscitation (CPR).

**Method:** Articles were searched on the international bases of scientific studies such as PubMed, PubMed Central, Medline, Cochrane Library, and Google Scholar. This review was carried out using meta-analysis and international guidelines.

**Results:** Four articles were included that met the inclusion criteria. It appears that the likelihood of a bystander implementing CPR is increased when supported by telephone instructions. Factors such as anxiety and knowledge affect the quality of CPR provided. Finally, short and encouraging instructions from the health services have greater effectiveness and a positive influence on the patient.

**Conclusion:** Telephone instructions are intended to provide CPR with the best possible quality and animate the resuscitator to perform the procedure correctly until a specialized health facility arrives. It appears that the likelihood of a bystander implementing CPR is increased when supported by telephone instructions. Regarding the limitations, there are differences between countries due to heterogeneous Health Service systems, level of education, and knowledge on the subject. In addition, there are barriers regarding the existing scientific knowledge, especially in Greece, where further investigation and awareness are necessary.

## Introduction

Cardiopulmonary resuscitation (CPR) refers to the emergency procedure that must be applied to any person suffering from cardiopulmonary arrest to keep them alive. Cardiopulmonary arrest is characterized by a sudden cessation of cardiac or respiratory function, or both, thereby causing insufficient tissue oxygenation, which, if not promptly reversed, is fatal.

In the causative factors of cardiopulmonary arrest, we distinguish the cardiac, respiratory, and hemodynamic causes, which respectively may come from pathological mechanisms or other non-pathological ones such as serious injury. In the main clinical manifestations of cardiopulmonary arrest, the following are noted: absence of pulse, absence of respiratory movements, possible obstruction of the airway, loss of level of consciousness, and cyanosis.^1,2,3,4^

Although CPR is an urgent and critical procedure, it consists of specific steps and skills, which must be applied in a specific sequence and with the best possible quality in order to be effective. Depending on the knowledge and skills of the person who will apply CPR and the place where the resuscitation will be attempted (community or health structure), there are different types of CPR, with the simplest and most common, Basic Life Support Support - BLS). Basic Life Support is performed by healthcare professionals and ordinary citizens who have been sensitized, trained, and certified in providing basic CPR.

Due to the importance of the immediate application of CPR in keeping the patient alive, some National First Aid Centers around the world provide the necessary instructions to the potential rescuer by telephone (Telecommunicator CPR - TCPR), who may not have been previously trained in CPR or is unable to recall and apply all the necessary steps correctly to perform CPR until the qualified health personnel of the First Aid services arrives at the incident.^5,6,7,8^

## Methods

For this systematic review, a search was performed in available databases (PubMed, Google Scholar, Embase, and Scopus) using a combination of the following search terms: CPR, Cardiac arrest, BLS, telephone CPR, first aid, tCPR.

Regarding the study design, case reports, cohort studies, cross-sectional studies, case series, and case-control studies were chosen. There were no language or other demographic restrictions.

Data from the eligible studies were extracted, including the name of the first author, region/country where the survey was conducted, study period, study design, sample size, outcomes or way/questionnaires which were used, and main findings concerning the efficiency of the telephone instructed CPR.

Criteria for inclusion and exclusion of the studies under evaluation were set. The following were used as inclusion criteria:

- Studies carried out in the last ten years (2012-2022).
- Articles with free access.
- Studies conducted in the human population.
- Studies conducted in an adult population.

The process of selecting and excluding articles is captured in **Figure 1**.

**Figure 1:**
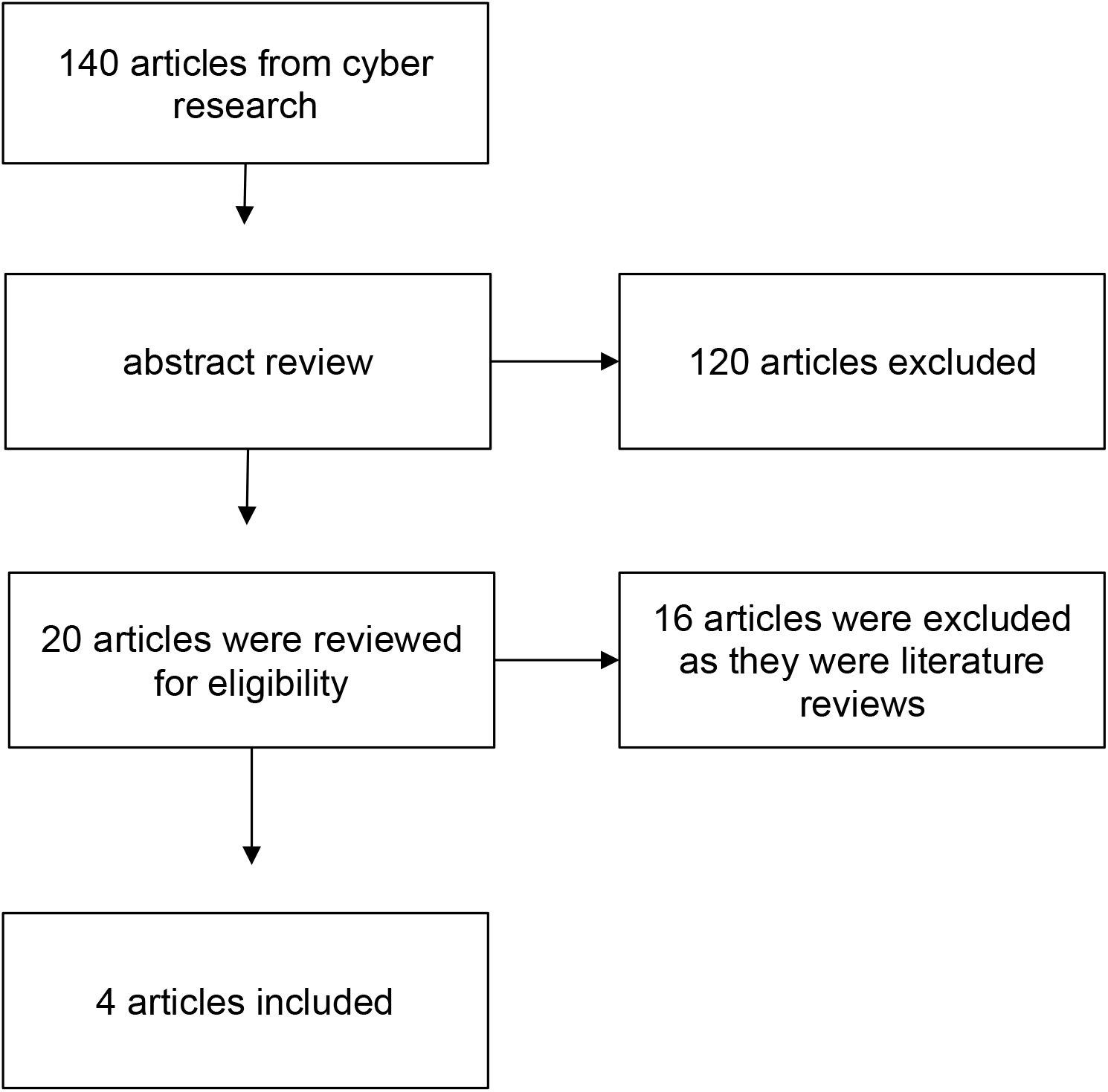
Flow chart of the systematic review.

Finally, four studies were included. These studies came from Washington (one study), Singapore (one study), Australia (one study), and Russia (one study). No quantitative synthesis of results was performed, but only a systematic review of studies was performed. Also, no assessment of the quality of the studies based on specific tools was attempted because the purpose of the specific research was descriptive.

## Results

### Lewis et al^9^

**Table 1.** Summary of the studies in the systematic review.

| Study | Country and<br>year of<br>publication | Type of<br>study | Aim | Population and<br>Methodology | Results |
| --- | --- | --- | --- | --- | --- |
| Lewis et al<sup>9</sup> | Washington, 2013 | Retrospective Cohort Study | The identification and characterization of the specific factors leading to non-recognition of cardiac arrest by bystanders | In 590 out-of-hospital cases of cardiorespiratory arrest during the year 2011, the correct identification of cardiac arrest and the provision of CPR were observed. | Correct recognition of cardiorespiratory arrest in 80% of cases. The mean arrest recognition time was 75 seconds and telephone-guided CPR was applied to 62% of patients and the mean CPR time was 176 seconds |
| Weng Kee Leong et al<sup>10</sup> | Singapore, 2021 | Cross-sectional study | Investigating the effectiveness of simplified telephone instructions in performing CPR | There were created 2 groups of application of different telephone instruction (press 100/min with 5cm depth n=282 Vs press hard and fast n= 24) | Compliance in the first group was 15.2% and in the second group 72.8%. In the first group the average time for the first compression was 34.36 seconds and in the second group 26.83. The need to explain the instruction was 60.4% in the first group and 81.5% in the second. |
| Marine Riou et al<sup>11</sup> | Perth, Australia, 2018 | Prospective cohort study | Investigating the effectiveness of telephone instructions from call center paramedics in motivating bystanders to initiate CPR. | Emergency calls (n=424) of identified cardiac arrest episodes were used. From these, the positive response of the attendants to the initiation of CPR was investigated in relation to the verbal choices of the call center rescuer. | CPR was performed in 85% of all calls. A greater positive response was observed in the groups where the rescuer gave a strict instruction ["We will do CPR (97%) and "We need to do CPR" (84%)]. Conversely, smaller percentages (43%) were noted when the telephone instruction was with a question ("Shall we perform CPR?") |
| Alexei Birkun et al<sup>12</sup> | Crimea, 2018 | Prospective blinded randomized control study | Evaluating the effectiveness of videotaped CPR-guided telephone instructions compared to simple call center telephone instructions. | 2 CPR execution teams were created: team 1, n=57 Guided by audio-visual material and group 2, n=52 guided by phone instructions only. In both groups, CPR performance was assessed based on a list of 13 parameters. | There were no major differences between the groups regarding the final score of the list ($5.6 \pm 2.2$ vs. $5.1 \pm 1.9$, $P > 0.05$). Group 1 had a shorter time to first compression and a greater total number of compressions than group 2. |

A retrospective cohort study published in 2013 investigated the identification and characterization of specific factors that led bystanders not to recognize cardiac arrest. In terms of methodology, the correct recognition of cardiac arrest and the provision of CPR was observed in 590 out-of-hospital cases of cardiorespiratory arrest in the year 2011. In the results, it is noted that the probability of applying CPR by a bystander doubled when it was guided by telephone by the respective health services. Although even in this case, i.e., the provision of assistance by the Health Professional at the call center, there is difficulty in correctly identifying the episode of cardiac arrest, in 80% of the calls, the existence of the arrest was successfully recognized, and therefore, the need for immediate application of CPR.

### Weng Kee Leong et al^10^

In a cross-sectional study carried out in Singapore, the effectiveness of the instruction “push fast and hard” (s.p. on the chest) compared to the instruction “press 100 times to a depth of 5 cm” was studied in terms of speed and quality of the application. There Two groups were created for the application of different telephone instructions, with group 1 applying the instruction “push 100 times per minute with a depth of 5 cm” (n=282) and group 2 applying the instruction “press hard and fast” (n= 24). As a result, the “press fast and hard” instruction outperformed the other one. In more detail, compliance in the first group was 15.2%, while in the second group, 72.8%. In the first group, the average time for the first compression was 34.36 seconds, and in the second group 26.83. Furthermore, the need to explain the instruction was 60.4% in the first group and 81.5% in the second. This highlights the need to use short and simple instructions so that they can be understood.

### Marine Riou et al^11^

This is a prospective cohort study in which it investigated the effectiveness of telephone instructions from call center paramedics in motivating bystanders to initiate CPR. There were used Emergency calls (n=424) performed by bystanders in cardiac arrest episodes, recognized by the call center rescuers. From these, the positive response of the attendants to the initiation of CPR was investigated in relation to the verbal options provided by the call center rescuer. CPR was performed in 85% of all calls. A greater positive response was observed in the groups where the rescuer gave strict instructions (“We will do CPR” or “We need to do CPR” with 97% and 84% positive responses, respectively). Conversely, smaller percentages (43%) were noted when the telephone instruction was with a question (“Shall we perform CPR?”)

### Alexei Birkun et al^12^

This is a prospective, blinded, randomized study conducted in Crimea in 2018, which evaluated the effectiveness of telephone instructions for CPR, guided by videotaped audio-visual material, compared to simple telephone instructions from a call center. In terms of implementation, 2 CPR performance groups were created. Group 1 (n=57) was guided by audio-visual material, and group 2 (n=52) was guided by telephone instructions only. In both groups, CPR performance was assessed based on a list of 13 parameters. In the results, there were no major differences between the groups in terms of the final score of the list (5.6±2.2 vs. 5.1±1.9, P>0.05). Group 1 had a shorter time until the first compression and a greater total number of compressions than Group 2. In summary, no technique appeared superior to the other in terms of depth and rate of compressions however, the need for further investigation was highlighted.

## Discussion

CPR is an urgent and critical procedure to keep the victim alive. However, it consists of specific steps and skills to be ultimately effective. Due to the importance of the immediate application of CPR in keeping the patient alive, some National First Aid Centers around the world provide the necessary instructions to the potential rescuer by telephone (Telecommunicator CPR - TCPR).^1,2,3^

It is noted that the probability of applying CPR by a bystander increases when it is guided by telephone by the respective health services. In the research of Lewis et al. and Marine Riou et al.,^9,11^ it is shown that the participation of the rescuer through the telephone, in combination with the motivation of the bystander using strict instructions, contributes to the increase of the positive response rates of the bystander and finally, application of CPR.

Regarding the quality of CPR when guided by telephone instructions, it depends on a series of factors such as the pre-existing knowledge and stress of the rescuer, the way the algorithm is transmitted and communicated by the health professional, the anatomy of the patient, and others. Regarding the transmission of the instructions for the implementation of the algorithm, these differ between the health services of each country. However, the research carried out in Singapore, it was studied the effectiveness of the directive “push fast and hard” (s.p. chest) compared to the directive “push 100 times to a depth of 5 cm” in terms of speed and quality of the application. In the results, the instruction “press fast and hard” prevailed over the second, while in 60% of the participants, it was necessary to explain the instructions to carry them out, highlighting the need to use short and simple instructions to understand them.

The research by Marine Riou et al.,^11^ its was emphasized the importance of promoting CPR by the healthcare professional to the rescuer caller, using phrases such as “We will apply CPR” and avoiding phrases such as “Do you want to apply CPR?”. When the first phrase was used, 97% of the callers responded positively, while in the second phrase, 84%. Thus, it emerges that the combination of short and “imperative” phrases is more effective in terms of the quality and speed of CPR application.^10,11^

Finally, it appears that the use of audio-visual media slightly, based on the research of Alexei Birkun et al., enhances the effectiveness of CPR compared to simple telephone instructions without, however, having more research data to draw valid conclusions.^12^

## Conclusions

Telephone instructions are intended to provide CPR with the best possible quality and animate the resuscitator to perform the procedure correctly until a specialized health facility arrives. It appears that the likelihood of a bystander implementing CPR is increased when supported by telephone instructions. Factors such as anxiety and knowledge affect the quality of CPR provided. Finally, short and encouraging instructions from the health services have greater effectiveness and a positive influence on the patient.

Regarding the limitations, there are differences between countries due to heterogeneous Health Service systems, level of education, and knowledge on the subject. In addition, literature references with a satisfactory sample are not observed, creating a gap in drawing conclusions and improving the care provided. Especially in Greece, there is no corresponding research that studies the quality of CPR provided by the general population using telephone instructions in combination with the observation of other demographic characteristics. The aim is to increase the awareness and information of healthcare workers and citizens regarding familiarity with the field of CPR for the best possible treatment of cardiopulmonary resuscitation cases in the community.

## Data Availability

All data produced in the present study are available upon reasonable request to the authors

